# Clinical outcomes and its determinants among neonates with neonatal sepsis admitted to selected Governmental hospitals in Addis Ababa, Ethiopia

**DOI:** 10.64898/2026.08.10.26360094

**Authors:** Rebuma Muleta Gutema, Galana Takele Namara

## Abstract

**Background:** Even though significant advances in diagnosis, treatment, and prevention strategies have been implemented, neonatal sepsis remains a common concern in clinical practice, especially in low-resource countries. It is one of the major causes of death during the first month of life. This study aimed to assess clinical outcomes and predictors of mortality among neonates with neonatal sepsis admitted to public hospitals in selected Hospitals in Addis Ababa, Ethiopia.

**Methods:** A hospital-based prospective cohort study design was conducted among 466 neonates admitted with neonatal sepsis from September 2024 to January 2025. All neonates who were admitted to selected Hospitals of Addis Ababa city after being clinically or laboratory-diagnosed with neonatal sepsis by the attending physician were included in the study. Data were entered into EpiData 4.2 and analyzed by SPSS version 26. Bivariate and multivariate Cox regression were used to identify the relationship between dependent and independent variables. Finally, variables with p-value ≤ 0.05 were taken as significant factors associated with poor clinical outcome.

**Results:** The study was conducted among 466 neonates admitted with neonatal sepsis. Of all neonates admitted with neonatal sepsis, 372 (79.8%) were discharged with good outcomes, and (20.2%) had a poor outcome/died. Duration of ruptured membrane being >12hr (AOR=7.02, % (CI: 1.85, 26.57), marital status /divorced (AOR=3.12, 95 % (CI: 1.67, 7.45),rural residence (AOR= 6.05, 95 % (CI: 2.03-16.53), assisted instrumental delivery (AOR= 5.99, 95 % (CI: 1.46-17.11), meconium-stained amniotic fluid ((AOR= 9.48, 95 % (CI: 0.49-18.61)), no initiate exclusive breast feed within one hour (AOR= 3.20, 95 % (CI: 0.90-7.52), chest in drawing (AOR= 5.81, 95 % (CI: 1.75-11.23) were significantly associated with neonatal mortality.

**Conclusion:** Neonatal mortality was moderately high. Meconium-stained amniotic fluid, prolonged duration of ruptured membrane (>12hr), Mode of delivery (instrumental delivery), and chest in drawing are among the predictors of neonatal mortality.

## INTRODUCTION

### Background

Neonatal sepsis refers to an infection involving the bloodstream in newborn infants less than 28 days old, as a result of a suspected or proven causative agent [1]. Neonatal sepsis is classified into two main categories: early onset neonatal sepsis (EONS) and late onset neonatal sepsis (LONS). Early onset appears within the first three days in most cases. Early-onset sepsis (EOS) is generally caused by the transmission of pathogens from the female genitourinary system to the newborn or the fetus [2]. Bacterial sepsis is considered to be the most common cause of neonatal mortality (deaths) in the first month of life [3]. The most common organisms in EONS are Group B Streptococcus (50%) and Escherichia coli (20%). Late onset neonatal sepsis in other way happens from 4 days of neonatal life and is mostly developed after delivery [4]. Late-onset neonatal sepsis (LOS) usually occurs via the transmission of pathogens from the surrounding environment after delivery, such as contact from healthcare workers or caregivers, late occurrence of vertically transmitted infection, and infants who undergo invasive procedures that disrupt the mucosa [5]. Around 70% LONS are due to gram-positive infections, such as Coagulase-negative staphylococci, Staphylococcus aureus, and Enterococcus [6].

Maternal factors that increase the risk of neonatal sepsis include chorioamnionitis, group B streptococcus colonization, preterm, and prolonged rupture of membranes greater than 18 hours [7]. The incidence of EOS has decreased since the 1990s due to the introduction of universal screening of group B streptococcus in pregnant women and intrapartum antibiotic prophylaxis [8]. However, rates of LOS have remained relatively the same. Although the availability of injectable antibiotics for infants with sepsis is increasing, deaths during the first month of life continue to rise in their proportion of total child mortality before 5 years of age [9]. Effective treatment of infants presenting with clinical signs of sepsis is a life-saving intervention, but the antimicrobial resistance that follows, especially as a result of broad-spectrum antibiotic use, is threatening future progress [10, 11].

The most common clinical manifestations of neonatal sepsis are altered behavior, muscle tone, feed refusal, feed intolerance including: vomiting, excessive gastric aspirates and abdominal distension, temperature instability, hypotension, poor perfusion with pallor and mottled skin, metabolic acidosis, tachycardia or Bradycardia, apnea, respiratory distress, grunting, cyanosis, irritability, lethargy, seizures, petechial, purpura, and bleeding. Therefore neonatal sepsis can be diagnosed if at least two of the above clinical manifestations feature and at least two of the following laboratory values are positive, such as, complete blood count with differential, blood culture, urine culture, chest radiography (if respiratory signs present), gram stain, lumbar puncture examination especially for late-onset sepsis [12-16].

Neonatal sepsis remains a major cause of neonatal morbidity and mortality. The World Health Organization estimated that there are approximately five million neonatal deaths per year of which 98 % occur in developing countries[17, 18]. In sub-Saharan Africa, neonatal sepsis contributed 17% of neonatal deaths and an estimated 49.6% of all under-five deaths in 2013 [19]. From those neonatal deaths, most of the deaths occurred in the first month, and from those who die in the first month, two-thirds of deaths occur in the first week, and of those who die in the first week, two-thirds die in the first 24 hours [20]. In other words, short-term and long-term complications occur among the surviving neonates with neonatal sepsis [21]. Reducing newborn and under-five mortality as low as 12/1000 and 25/1000, respectively, is one of the global strategies of the WHO in African countries by 2030 to achieve the Sustainable Development Goal [22]. This could be achieved through better prevention and management of severe infections in newborn [23, 24]. Neonatal sepsis is the most common cause of neonatal death, contributing 26 % next to preterm in Ethiopia [25]. According to the Ethiopian demographic and health survey report, there has been a slight increase in neonatal mortality from 29 deaths per 1,000 live births in 2016 to 33 deaths per 1,000 live births in 2019 [26]. Despite this massive contribution to neonatal death still the problem related to neonatal sepsis treatment and contributing factors were not well studied in general and in the study area in particular. Therefore, this study aimed to assess clinical outcome and predictors of neonatal sepsis among neonates admitted to public hospitals of Addis Ababa, Ethiopia.

## METHODS

### Study Design, Area, and Period

A longitudinal prospective cohort study was carried out at selected public hospitals in Addis Ababa from September 2024 to January 2025. Addis Ababa is the capital city of Ethiopia, and a Seat for the African Union, and the United Nations Economic Commission for Africa. The area of the city was 527 square kilometers and had 12 sub-cities. According to a population projection for 2023, the city has an estimated population of 6.1 million [27]. The city has 13 public Hospitals, and all of them have neonatal intensive care unit. Among these 7 were under the Addis Ababa Health Bureau, 5 were under the Ministry of Health, and 1 was under Addis Ababa University[28]. The study was conducted in four selected Addis Ababa public Hospitals. These selected Hospitals are: Zewuditu Hospital, St. Peter Specialized Hospital, Tikur Anbessa Specialized Hospital, and Yekatit 12 Hospital Medical College. In Zewuditu Hospital: The number of admitted neonates varies from time to time; the average annual admission rate was 3,325. The NICU had a 48-bed capacity. It has radiant warmers to keep the room warm and 12 incubators for premature neonates. In St Peter Specialized Hospital, the number of admitted neonates on average was 2600. The NICU had a 34-bed capacity. In Tikur Anbessa Specialized Hospital; the average annual admission rate neonatal sepsis was 2740. The Unit had a 41-bed capacity (50). In Yekatit 12 Hospital Medical College: the average annual admission rate was 3000. The intensive care unit had a 45-bed capacity.

### Sample size and sampling procedure

The sample size was determined by using a single population proportion formula by considering the following assumptions: 95% CI, 5% marginal error, and by taking culture-proven neonatal sepsis, approximately 24.4 % in the previous study in Ethiopia [29]. Using a design effect of 1.5 to quantify the sample and 10% for non-response, the final sample size was 466.

### Operational definitions of the variables

**Neonate:** is a baby from birth until 28 days of life.

**Neonatal sepsis:** an infection involving the bloodstream in newborn infants less than 28 days old with the presence of at least two risk factors and/or clinical features of bacterial infections plus at least two laboratory results which are suggestive for neonatal sepsis or neonates who are diagnosed as sepsis by attending physician and fulfill sepsis criteria.

**Early onset of neonatal sepsis** is sepsis/infection occurring from birth to three days of life.

**Late onset of neonatal sepsis:** sepsis/infection happening from 4 to 28 days of age.

**Clinical outcomes:** a neonatal condition in health, function, or quality of life resulting from medical interventions, serving as key indicators of treatment effectiveness, safety, survival rates, and symptom improvement.

**Improved:** If the neonate improves after completing the treatment of sepsis without any complications.

**Not Improved:** If the neonate does not improve after completing the treatment, deteriorates, is referred to, or dies.

### Study variables

#### Dependent variables: clinical outcome of neonatal sepsis

Independent variables: Socio-demographic characteristics such as (Maternal age, maternal marital status, Residence, monthly income and Educational status of mother), Maternal factors like (Parity, History of UTI, Foul smelling fluid/vaginal discharge, Antenatal care, Duration of labor), Neonatal Factors such as (Age of the neonate, Sex of the neonate, APGAR score, Resuscitation at birth, Gestational age), Clinical presentation of neonatal sepsis: (Tachypnea, Poor feeding, and Fever), Diagnostic/laboratory results of neonates (CBC, protein, glucose and Blood culture), and Antimicrobial use in neonatal sepsis.

#### Data Collection tool and Procedure

Data were collected by trained data collectors using a structured questionnaire and checklist. Both primary and secondary data were used. Data quality was assured by carefully designing the data extraction tools and training both the data collectors and supervisors. Moreover, pretesting among 5% of the sample size outside of the study area was made to assess for its completeness, clarity, length, skip patterns, and correctness of filled questionnaires.

In this study, neonatal sepsis was diagnosed as either of the clinical manifestations (changes in temperature, feeding problem, fussiness, lack of energy, high-pitched cry, yellow, blue, or pale skin, bruising or bleeding, cool, clammy skin, skin rashes, fast breathing, breathing problems, or periods of no breathing, vomiting, and diarrhea) and at least one positive laboratory test for a bacterial pathogen (It could be positive bacterial culture result/polymerase chain reaction (PCR)/gram-staining/latex agglutination tests/antigen-antibody detection for bacteria).

### Data analysis procedure

Data were cleaned, coded, and entered into Epi Data version 4.6 software, and then exported to SPSS version 26 for analysis. Then, exploratory data analysis was carried out to check the levels of missing values, multi-collinearity, and proportionality of hazards over time. To identify associations between dependent and independent variables, bivariate analysis was conducted.

Crude odds ratio and adjusted hazard ratio, at 95% CI and p-value <0.05, were used to assess the strength of association and statistical significance. Variables significant at the P < 0.05 level in the bivariate analysis were considered and included in the final Cox-regression analysis, to identify independent predictors of mortality.

## Result

In this study, 466 neonates with their index mothers were enrolled. The minimum and maximum ages of the neonates were one day and twenty-six days, respectively, with a mean of 4:54 ± 6:02 days. Nearly two-thirds (65.7%) of the neonates had an age of less than three days. Among the neonates who died with neonatal sepsis, 66(70.12%) of the deaths occurred within the first three days (72 hours). Regarding the sex of the neonates with neonatal sepsis, 246 (52.8%) were males, and among them, 48(51.06%) died, which makes nearly equal among males and females. The minimum age of the mother was 20 years, and the maximum age was 40 years, with a mean of 24.18 ± 3:29 years. In considering the mortality, 41(44%) neonates died among mothers aged > 30 years. A majority of 385(82.6%) of the mothers were got married and about 297 (63.7%) mothers’ were rural residents. Regarding the maternal educational status, 124 (26.6%) had no formal education, while 108 (23.2%), 176 (37.8%), and 58 (12.4%) accounted for Primary education, secondary education, College and above, respectively. Regarding the maternal occupational status, most of them, 205 (44.0%), were housewives, while 30(6.4 %) were private employees (Table 1)

**Table 1:** Socio-Demographic Characteristics of the Neonate with their Mothers Admitted to public hospitals, Addis Ababa, Ethiopia 2024 (n = 466).

| Variables | Category | Frequency | Percent |
| --- | --- | --- | --- |
| **Age of the newborn at admission** | Birth-3 days | 306 | 65.7 |
|  | 4–28 days | 160 | 34.3 |
| **Sex of the newborn** | Male | 246 | 52.8 |
|  | Female | 220 | 47.2 |
| **Mother's age** | 18–23 | 187 | 40.12 |
|  | 24–29 | 246 | 52.78 |
|  | >30 | 33 | 7.08 |
| **Marital status** | Married | 385 | 82.6 |
|  | Single | 9 | 1.9 |
|  | widowed | 21 | 4.5 |
|  | Divorced | 51 | 10.9 |
| **Residence** | Urban | 297 | 63.7 |
|  | Rural | 169 | 36.3 |
| **Maternal education** | No formal education | 124 | 26.6 |
|  | Primary education | 108 | 23.2 |
|  | Secondary education | 176 | 37.8 |
|  | college and above | 58 | 12.4 |
| **Maternal Occupation status** | Housewife | 205 | 44.0 |
|  | Governmental employee | 135 | 29.0 |
|  | Merchant | 96 | 20.6 |
|  | private employee | 30 | 6.4 |
| **Father educational status** | No formal education | 8 | 1.6 |
|  | Primary education | 67 | 14.4 |
|  | Secondary education | 234 | 50.2 |
|  | college and above | 157 | 33.7 |
| **Economic status (monthly income)** | < 2000 | 94 | 20.2 |
|  | 2001-3000 | 43 | 9.2 |
|  | 3001-4000 | 129 | 27.7 |
|  | 4001-5000 | 97 | 20.8 |
|  | >5000 | 103 | 22.1 |

### Maternal Related Factors

More than half of the women, 270(57.9%), were primigravida. Majority 404(93.2%) of the women had ANC at least one contact/visit,266(57.1 %) women had a history of Chorioamnionitis during current birth, of these 62(65.9%) of neonate died,166(35.6%) of woman developed PROM/PPROM, among these110(66.26%) have < 12 hours duration of rupture of membrane. Regarding place of delivery, 361(77.46 %) of the women delivered their newborn in a health institution, 290 (62.3%) women delivered spontaneously,190(40.8%)of the women developed meconium-stained amniotic fluid, and 103(22.2%) had a history of bleeding during the current pregnancy (Table 2).

**Table 2:**
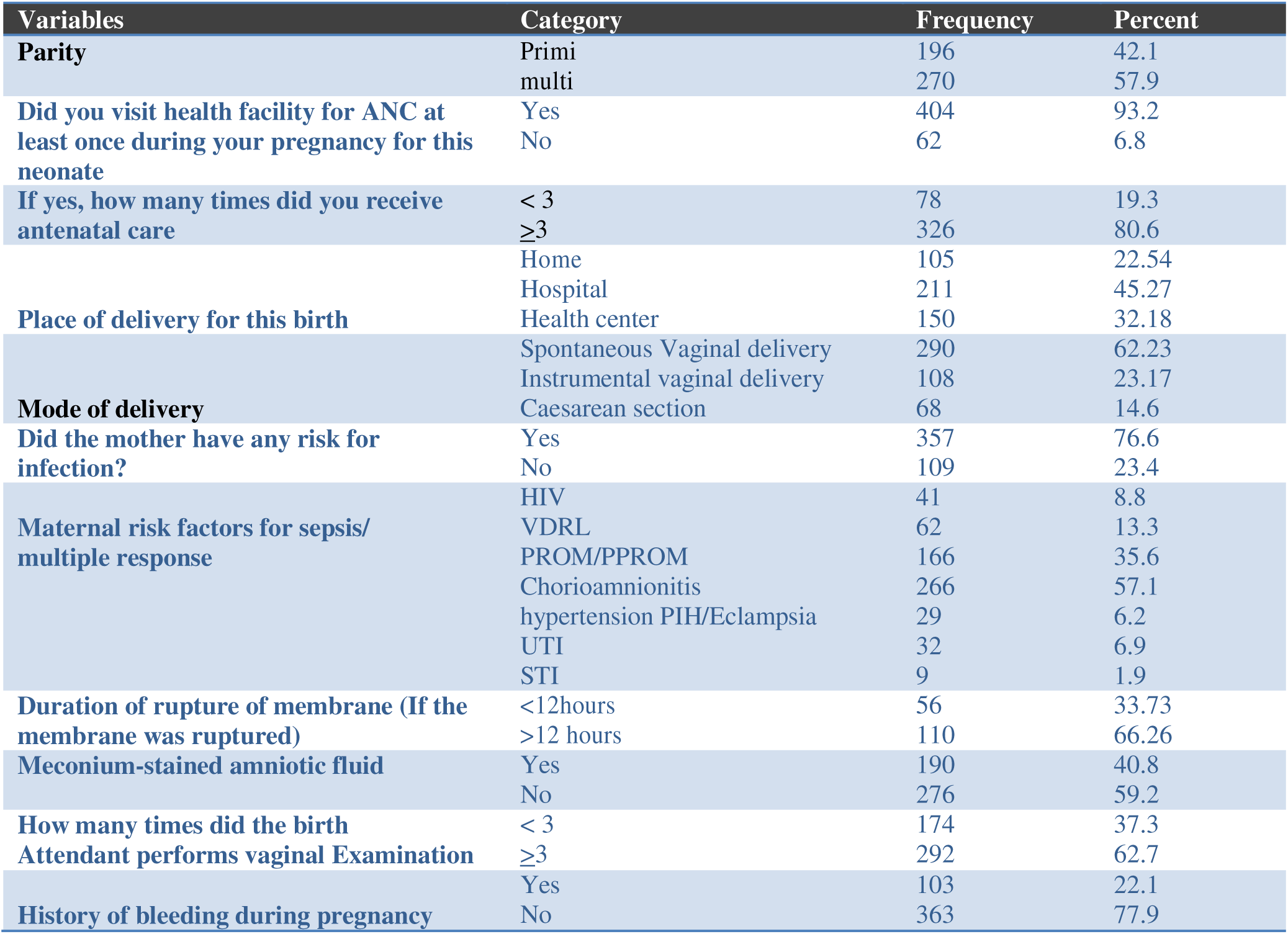
Maternal Related Characteristics for Clinical outcome of Neonatal Sepsis Admitted to public hospitals, Addis Ababa, Ethiopia 2024 (n = 466).

### Neonatal Characteristics

Regarding gestational age at birth, 81(17.4%) was preterm while 7(1.5%) were post term, 74(15.91%) was weight<2500gm, and 196 (42.1%) neonates had 1stminute Apgar score of 0-3, among which majority of the neonate 76 (80.85%) died. Among the admitted neonate majority, 323(69.3%) of the neonates have a history of risk of infection, like resuscitation at birth 201(43.1%) of these 63(13.5%) dead, followed by Prematurity 206(44.2%) of which only 20(4.3%) have poor outcome. Of the total participants, 416(89.3%) women initiated exclusive breastfeeding within one hour(Table 3).

**Table 3:**
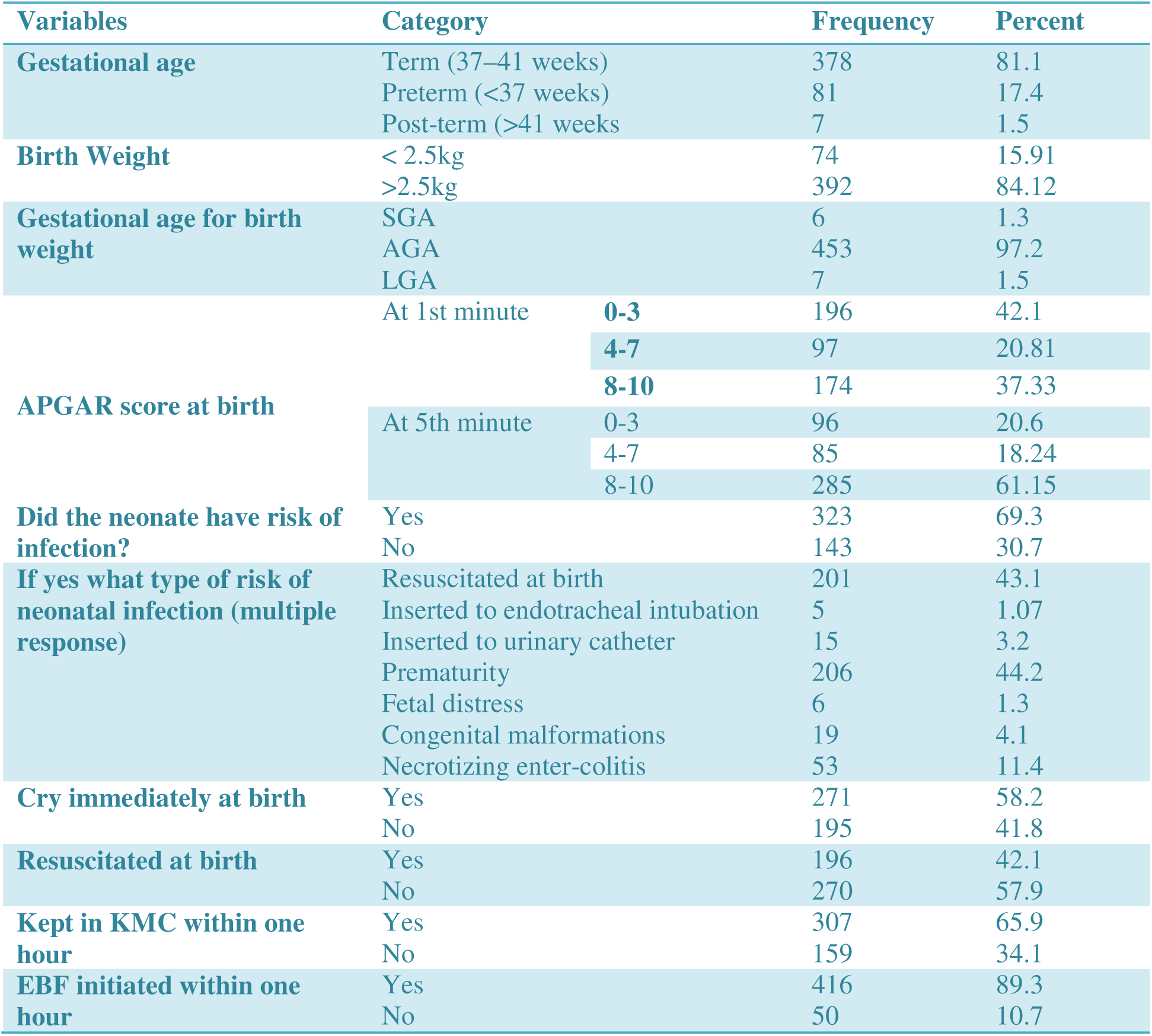
Neonatal Related Characteristics for Clinical outcome of Neonatal Sepsis Patients Admitted to public hospitals, Addis Ababa, Ethiopia2024 (n = 466).

### Clinical Parameters and Laboratory Findings

During admission time, about 366(78.5%), 446 (95.7%), 399(85.6%), and 431 (92.5%) of the neonates had respiratory bradypnea or tachypnea, Pulse rate bradycardia or tachycardia, nasal flaring, and fever (>37.5), respectively.

Of all neonatal sepsis patients, the following laboratory findings were also performed in addition to clinical presentation. About 393(84.3%) were diagnosed by CBC, 172(36.9%) were diagnosed with culture, 403(86.5%) with gram stain, and 347 (74.5%) with a CSF test for diagnosing neonatal sepsis (Table 4)

**Table 4:** Clinical Presentation and Vital Sign of Neonatal Sepsis Patients Admitted to public hospitals, Addis Ababa, Ethiopia2024 (n = 466).

| Variables | Category | Frequency | Percent |
| --- | --- | --- | --- |
| Respiratoryrate(bradypniaor tachypnea)(Count full minute) | Yes | 366 | 78.5 |
|  | No | 100 | 21.5 |
| Severe chest in drawing | Yes | 197 | 42.3 |
|  | No | 269 | 57.7 |
| Nasal flaring | Yes | 399 | 85.6 |
|  | No | 67 | 14.4 |
| Grunting | Yes | 80 | 17.2 |
|  | No | 386 | 82.8 |
| Vomiting | Yes | 188 | 40.3 |
|  | No | 278 | 59.7 |
| Pulse rate (bradycardia or tachycardia) in beat/minute | Yes | 446 | 95.7 |
|  | No | 20 | 4.3 |
| Lethargic or unconscious | Yes | 413 | 88.6 |
|  | No | 53 | 11.4 |
| Reduced movements (activity) | Yes | 399 | 85.6 |
|  | No | 67 | 14.4 |
| Unable to feed (failure to suck) | Yes | 372 | 79.8 |
|  | No | 94 | 20.2 |
| Abdominal distention | Yes | 372 | 79.8 |
|  | No | 94 | 20.2 |
| Fever (>37.5) | Yes | 431 | 92.5 |
|  | No | 35 | 7.5 |
| Type of sepsis | Earlyonset | 305 | 65.5 |
|  | Late onset | 161 | 34.5 |
| CBC results had drop/ raise (Platelet, Neutrophil, WBC, HCT Abnormal (raise/drop) (if done) | Normal | 53 | 13.83 |
|  | Abnormal (raise/drop | 330 | 86.16 |
| test result ofC reactive protein (CRP) (if done) | Reactive | 98 | 42.6 |
|  | Non-reactive | 132 | 57.3 |
| Blood culture result (if done) | Normal | 61 | 22.93 |
|  | Abnormal (raise/drop) | 172 | 36.9 |
| If done and abnormal Identified causative organism | Escherichia coli | 52 | 30.23 |
|  | Group B streptococci | 92 | 53.48 |
|  | Listeria monocytogenes | 28 | 16.27 |
| Gram stain done | Positive | 103 | 81.75 |
|  | Negative | 23 | 18.25 |

#### Treatment received and outcome

Most of 254(54.4%) neonates were received treatment with ampicillin plus gentamycin, 80 (17.16%) with a combination of ampicillin, gentamycin, and ceftriaxone; 105 (22.5%) with a combination of Ampicillin, gentamycin, and Vancomycin, and 24(5.15%) with a combination of gentamycin and ceftriaxone.

The clinical outcome of the study was a poor outcome, and a good outcome. Of all neonatal sepsis patients admitted to the NICU, 372 (79.8%) of the neonates were discharged with good outcomes, and 94 (20.2%) had a poor outcome/died (Table 5)

**Table 5:** Treatments received for the treatment of neonatal sepsis by the neonates Admitted to public hospitals, Addis Ababa, Ethiopia2024 (n = 466).

| Variables | Category | Frequency | Percent |
| --- | --- | --- | --- |
| Ampicillin+ Gentamicin | Yes | 254 | 54.4 |
|  | For how many days | 7 |  |
| Cefotaxime + Amikacin | Yes | 3 | 0.6 |
|  | For how many days | 21 |  |
| Ampicillin + Gentamicin + cefotaxime | Yes | 80 | 17.16 |
|  | For how many days | 14 |  |
| Ampicillin + genta + Vancomycin | Yes | 105 | 22.5 |
|  | For how many days | 10 |  |
| Gentamycin + ceftriaxone | Yes | 24 | 5.15 |
|  | For how many days | 10 |  |
| Was neonate on oxygen? | Yes | 317 | 68.02 |
|  | No | 149 | 31.94 |
| If yes what was the method of oxygen administration? (multiple response) | Intranasal catheter (Nasal cannula) | 73 | 15.7 |
|  | Nasal prong (3 liter) | 140 | 30.0 |
|  | Nasal CPAP (> 4 liter) | 89 | 19.1 |
|  | Mask | 15 | 3.2 |
| Did the neonate have NG feeding tube inserted? | Yes | 174 | 37.3 |
|  | No | 292 | 62.7 |
| Clinical outcome | Good | 372 | 79.8 |
|  | Poor | 94 | 20.2 |

### Bivariate and Multivariable Cox Regression Analysis Results for Factors Associated with Mortality

Binary and multiple Cox regression analyses were used to investigate the association of independent variables with the dependent variable. In Bivariate Cox regression, duration of ruptured membrane, marital status, residence, mode of delivery, number of births, meconium-stained amniotic fluid, vomiting, EBF initiated within one hour, type of causative agent, chest in drawing, and abdominal distention were significantly associated with mortality for neonatal sepsis patients. On multivariate analysis, history of duration of ruptured membrane > 12 hours, marital status, residence, mode of delivery, meconium-stained amniotic fluid, EBF initiated within one hour, type of causative agent, and chest in drawing were among independent predictors of mortality associated with neonatal sepsis. Those neonates born to mothers who had a history of duration of ruptured membrane > 12 hours (AOR=7.02, 95 % CI: 1.85, 26.57) increased the risk of death by seven times compared to those whose duration is < 12 hours. Neonates from mothers of divorced marriages (AOR=3.12, 95 % CI: 1.67, 7.45) were three times more likely to die or have decreased survival compared to those neonates with mothers of married and living together. Neonates whose mothers reside in rural areas (AOR= 6.05, 95 % (CI: 2.03-16.53)) were six times more likely to die compared to those who are living in urban areas. Neonates whose mode of delivery was assisted Instrumental delivery (AOR=5.99, 95 % (CI: 1.46-17.11)) were nearly six times more likely to die compared to those who delivered spontaneously. Those neonates born to mothers who developed meconium-stained amniotic fluid (AOR= 9.48, 95 % (CI: 0.49-18.61)) were nine times more likely to die compared to those who did not develop it. Neonates whose mothers do not initiate exclusive breastfeeding within one hour (AOR= 3.20, 95 % (CI: 0.90-7.52) were three times more likely to die compared to those who do it. Neonates who develop chest in drawing (AOR= 5.81, 95 % (CI: 1.75-11.23) whose mothers reside in rural areas were six times more likely to die compared to those who live in urban areas (Table 6).

**Table 6:**
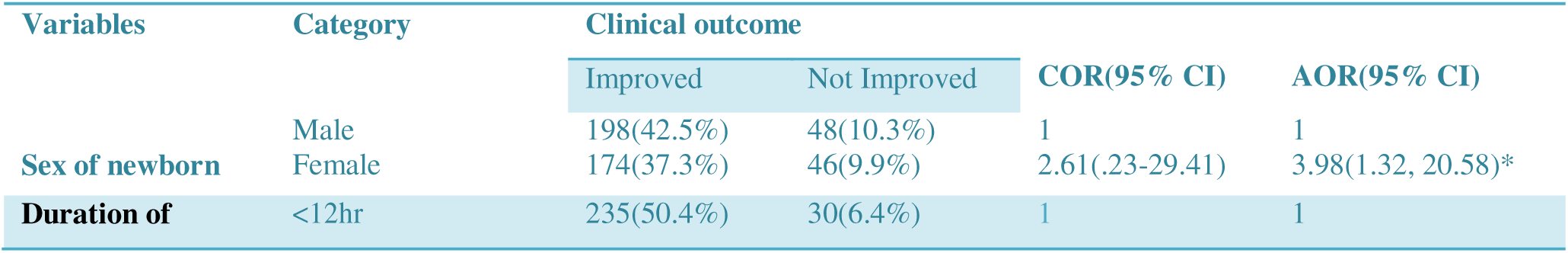
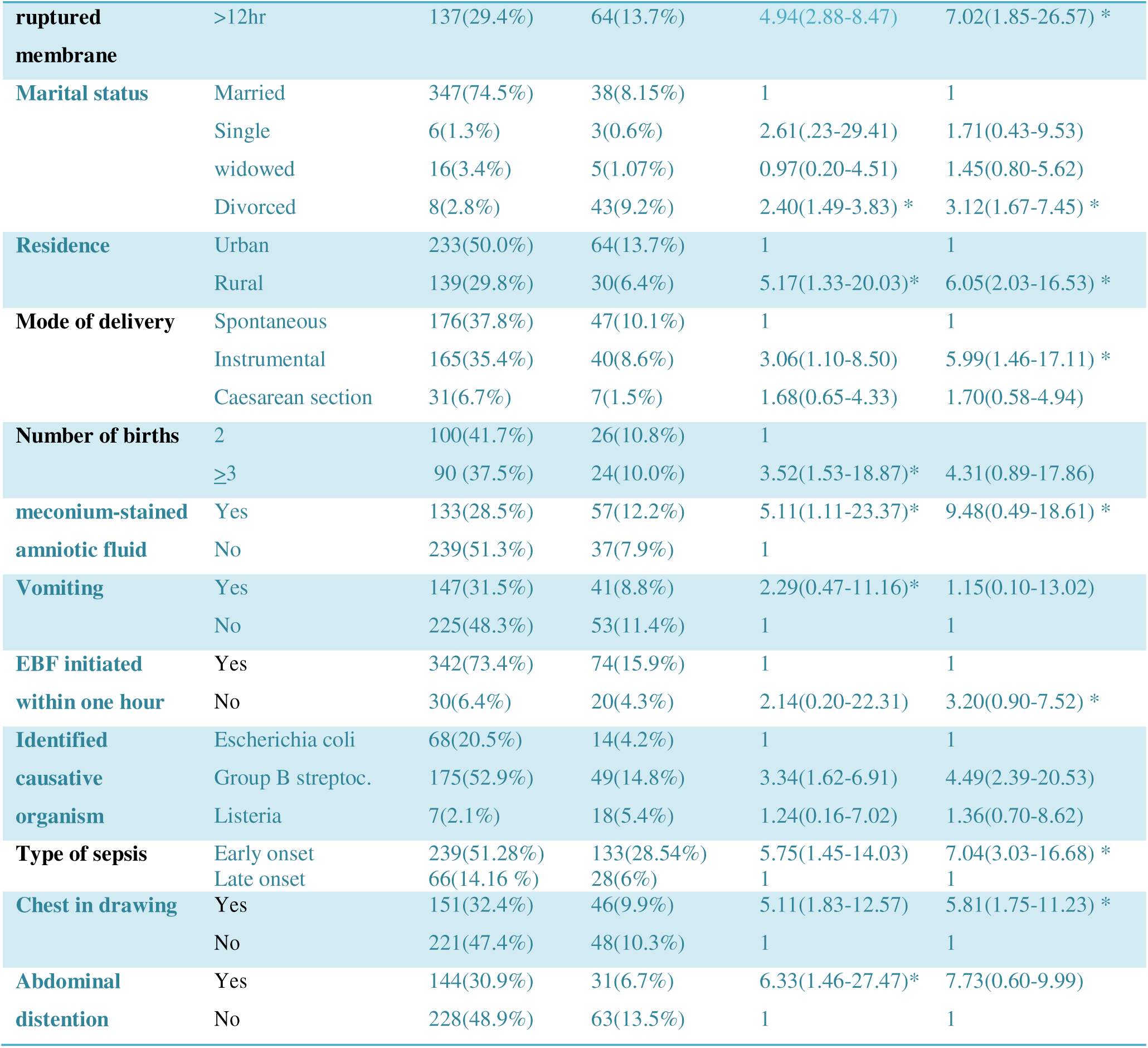
Cox-Regression Analysis for Factors Associated with clinical outcome Admitted to public hospitals, Addis Ababa, Ethiopia2024 (n = 466).

#### Survival Graph

The Log Rank (Mantel-Cox) (p=0.003), Breslow (p=008), and Tarone-Ware (p=0.023) indicate a significant difference in survival distributions between types of sepsis, suggesting that the type of sepsis affects survival time. On average, early-onset sepsis patients have a mean survival time of 4.81 ± 0.207 days (95% CI, 4.409–5.21) while it was 10.77± 0.293 days (95% CI, 10.10–11.24) for late-onset neonatal sepsis patients. This indicates that neonates with late-onset sepsis, on average, tend to survive longer compared to neonates with early onset. Early-onset neonatal sepsis [P = 0.003, AHR = 7.24, 95% CI: (3.03-16.68)] was 7 times more likely to cause death or decrease survival when compared to late-onset neonatal sepsis (Figure 1)

**Figure 1:**
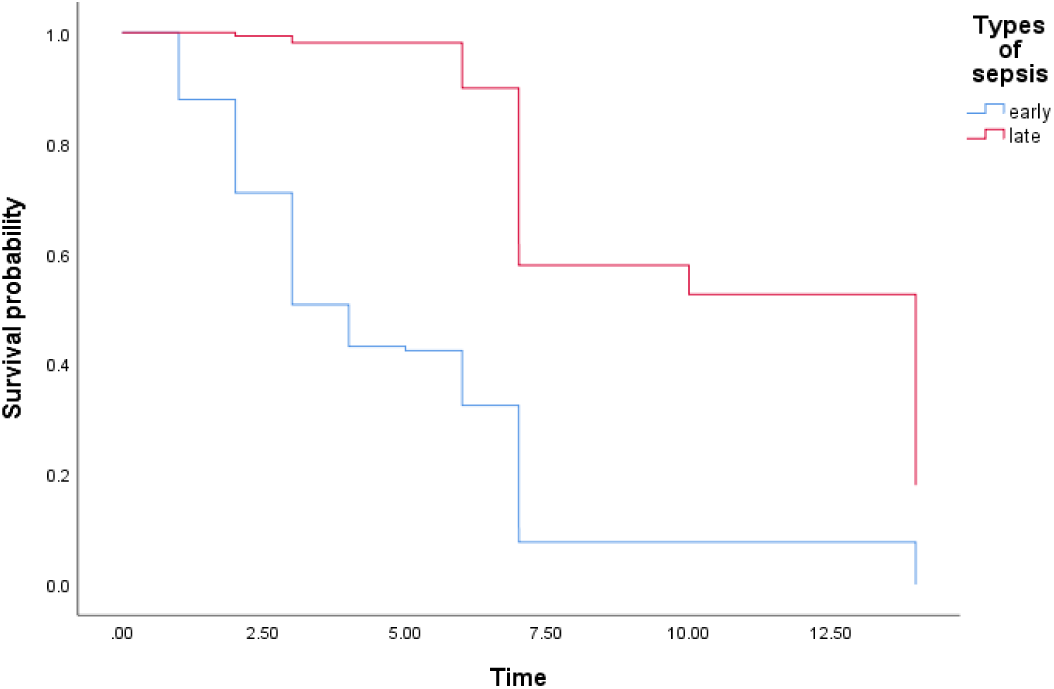
Kaplan–Meier estimation of survival based on types of sepsis among neonatal sepsis patients admitted to selected public hospitals in Addis Ababa 2024 (n = 466).

The Log Rank (Mantel-Cox) (p=0.01), Breslow (p=0.02), and Tarone-Ware (p=0.03) indicate a significant difference in survival distributions between sex of neonates, suggesting that sex of neonates affects survival time On average, female neonate have a mean survival time of 7.27 ± 0.50 days (95% CI, 6.27–8.27) while it was 10.06 ± 0.48 days (95% CI, 9.06–10.95) for male neonatal sepsis patients. This indicates that males, on average, tend to survive longer compared to female neonates with neonatal sepsis. Male neonate [P = 0.000, AHR = 3.98, 95% CI: (1.32, 20.58)] was 3 times more likely to cause death or decrease survival compared to female neonate with neonatal sepsis (Figure 2).

**Figure 2:**
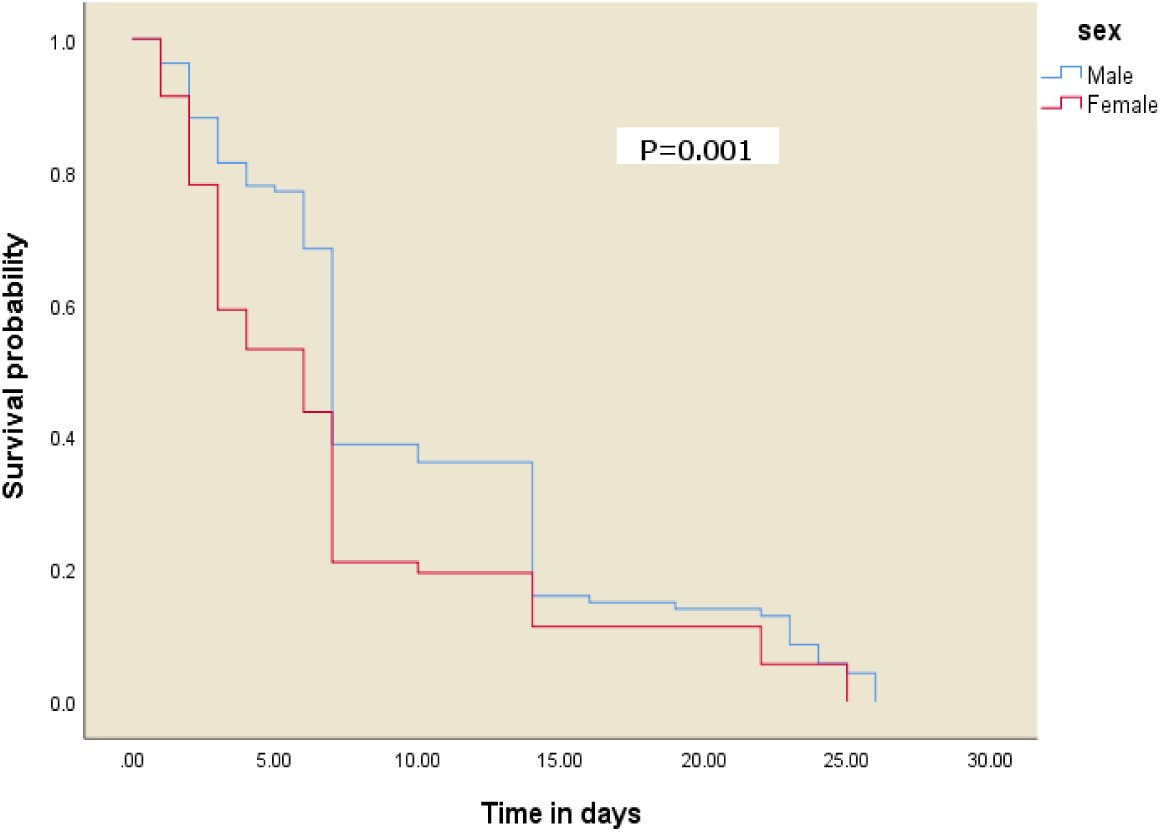
Kaplan–Meier estimation of survival based on sex among neonatal sepsis patients admitted to public hospitals of Addis Ababa 2024 (n = 466).

## Discussion

This study assessed clinical outcome and associated factors among neonatal sepsis patients admitted to Addis Ababa public Hospitals, Ethiopia. Having a prolonged duration of rupture of membrane, marital status, residence, mode of delivery, number of births, meconium-stained amniotic fluid, EBF initiated within one hour, identified causative organism, and Chest in drawing were statistically significant predictors of mortality. The overall neonatal mortality rate in this study was 20.2%. This finding is higher than a retrospective study done in the NICU of Manipal Teaching Hospital, Nepal (10%), in Mizan Tepi University Teaching Hospital, Ethiopia (14.7%), Hawassa University Comprehensive Specialized Hospital, Ethiopia(14.2%)and lower compared with studies conducted in Addis Ababa (24.4%), and Dubti General hospital, Ethiopia (27.2%) [6, 29-32]. These differences in mortality rate in neonatal sepsis among different countries may be explained by many factors such as socioeconomic, geographical, and racial factors, advanced care with advanced materials, use of ventilators, different strains of microorganisms, and use of different antibiotics.

This study revealed that neonates born to mothers whose rupture of the membrane was more than 12 hours before delivery have 7 times higher hazard of neonatal mortality than women whose membrane rupture was within 12 hours of delivery. This is similar to the study conducted in Jimma specialized hospital, in which those more than 12 hours have 7.74 times higher hazard of neonatal death, and Arba Minch General Hospital, which has 2.6 times higher hazard of neonatal mortality, respectively [33, 34]. This might be because, as the time of membrane rupture prolongs before delivery, ascending agents are the risk for sepsis, asphyxia, pulmonary hyperplasia, and preterm labor [35].

Neonates from women of divorced, are 3 times more likely to die or have decreased survival compared to those neonates with a mother of married and living with her husband. This might be because neonates with single parents may get less care, economic deficit, and nutritional problems.

Neonates whose mothers live in rural areas were 6 times more likely to die than those living in urban areas. This result differs in comparison to other studies reported at Jimma University Medical Center, Ethiopia [36]. The possible explanation might be due to the difference in geographical area, nutrition, availability of health care facilities nearby, sanitation, educational status, and health education coverage is better in urban than rural areas.

Neonates whose modes of delivery by assisted instrumental delivery were nearly six times more likely to die compared to those who were born spontaneously. This finding is supported by a study conducted at tertiary care hospitals in Sri Lanka, public hospitals of Sidama region, Southern Ethiopia, Hawassa University Comprehensive Specialized Hospital, and Adare General Hospital in Hawassa City, Ethiopia, respectively. This might be because, when the instrument is used, it may cause laceration of the newborn’s body, making it exposed to infection, as well as easily breakable neonatal mucous membrane, which can serve as a route of entry for infectious agents from contaminated equipment and environment [37-39].

Those neonates born to mothers who developed meconium-stained amniotic fluid were nine times more likely to die compared to those who did not develop it. This result is in line with the study conducted on neonatal sepsis at Public Hospitals in Jimma [40]. This might be due to the risk of meconium aspiration, which leads to neonatal infection and meconium aspiration syndrome.

The hazard of mortality among neonates with neonatal sepsis was 3 times higher if they did not initiate exclusive breastfeeding within one hour as compared with those who initiated within one hour (AHR: 3.20; 95% CI: 0.90-7.52). This finding is similar to the research conducted in Arba Minch General Hospital, Sawla General Hospital, and Chencha District Hospital [41]. This might be due to breastfeeding controlling the influence on the initial exposure of the newborn intestinal mucosa to microbes. Hence, limits disease-causing agents’ effect through the mucosa of the gut. In addition, the many defense factors of the mother’s milk include substantial amounts of secretory immunoglobulin A (SIgA) antibodies produced by lymphocytes that have migrated from the mother’s gut to the mammary glands. Therefore, the antibodies (secretory immune globulins) were commonly directed against the maternal past and recent microflora of the gut. In other words, the immediate initiation of exclusive breastfeeding acts as the initial point for a continuous maternal and newborn care that can have long-lasting effects on health and development, and provides adequate nutrition at an appropriate time, immunological value from first milk (colostrum), which prevents hypothermia and hypoglycemia [42,43].

## Strength and Limitation of the Study

This study includes higher sample size of neonatal sepsis diagnosed with advanced laboratory investigation (C reactive protein, culture, gram stain etc.) and experienced Physician compared to the others.

## Conclusion and Recommendation

This study indicated that mortalities among neonates with neonatal sepsis were slightly high compared to the national level. The majority (65.5%) of the neonates had early onset of sepsis. The result of this finding indicates sex, meconium-stained amniotic fluid, prolonged duration of ruptured membrane (>12hr), Mode of delivery (instrumental delivery), type of sepsis and neonates who has developed chest in drawing are among the predictive of neonatal mortality. Advanced and effective treatment was important for management of neonatal sepsis in this setup. Ampicillin and gentamicin combination was administered for majority of neonates with neonatal sepsis in this study. It showed all concerned body need to work hard for quality care and reduce neonatal death.

## Ethical Approval

This study was approved by Ethical review Committee (IRC) of Addis Ababa University with the number of AAU/R/E/A/05/405/2024. Informed consent was taken from each respondent before data collection of the study. Interviews were conducted in privacy, and participants were assured of confidentiality. This study was carried out in accordance to the pertinent ethical guidelines and regulations.

## Consent for publication

The study constitutes a low or no more than a minimal risk to the study participants. Also, the study did not involve any invasive procedures. Accordingly, after the objective of the study was explained, verbal consent was obtained from the parents or the guardian of the study participants. Moreover, the confidentiality of information was guaranteed by using code numbers rather than personal identifiers and by keeping the data locked.

## Data availability statement

The data that support the findings of this study will be made available from the corresponding authors, without undue reservation upon reasonable request.

## CRediT Author contributions statement

The Author involved in the conception, designing, analysis, and interpretation of the data. **Rebuma Muleta**; Conceptualized and designed the study, control and supervised the data collection, analyze the data and develop the manuscript, edited the data, and reviewed the manuscript. **Galana Takele**: Methodology, Writing – review & editing, Supervision, Validation. Finally, we agreed and approve of this article to be published in scientific journals and agreed to be responsible for all aspects of the work.

## Acknowledgments

We acknowledge all data collectors, supervisors, and respondents for their valuable hard work and responses for the success of this study. Finally, we sincerely thanks to our peers and staff for their valuable comments and proposition.

## Abbreviations

ANC: Antenatal care
AOR: Adjusted Odds Ratio
CI: Confidence interval
COR: Crude Odds Ratio
EONS: Early Onset Neonatal Sepsis
GBS: Group B streptoccoccus
LBW: Low birth weight
LONS: Late onset neonatal sepsis
MSAF: Meconium stained amniotic fluid
NICU: Neonatal intensive care unit
NMR: Neonatal mortality rate
OPD: Outpatient department
SPSS: Statistical Package for Social Science
TTBA: Traditional trained birth attendant
UTI: Urinary tract infection
WHO: World Health Organization

